# Relative Virulence of SARS-CoV-2 Among Vaccinated and Unvaccinated Individuals Hospitalized with SARS-CoV-2

**DOI:** 10.1101/2022.02.15.22271016

**Authors:** Alicia A. Grima, Kiera R. Murison, Alison E. Simmons, Ashleigh R. Tuite, David N. Fisman

## Abstract

**Background:** The rapid development of safe and effective vaccines against the SARS-CoV-2 virus has been a singular scientific achievement. Confounding due to health seeking behaviours and differential testing by vaccination status may bias analyses towards an apparent increase in infection severity following vaccination. We sought to determine whether risks of intensive care unit (ICU) admission and death were diminished significantly by vaccination, even in individuals for whom vaccination failed to prevent hospitalization.

**Methods:** We used data from Ontario, Canada’s Case and Contact Management database, merged to a provincial vaccination dataset (COVaxON) to create a time-matched cohort of individuals who were hospitalized with SARS-CoV-2 infection. Each vaccinated individual was matched to up to five unvaccinated individuals based on test date of positive SARS-CoV-2 infection. Risk of ICU admission and death were evaluated using multivariable conditional logistic regression. Unmatched exploratory analyses were performed to identify sources of heterogeneity in vaccine effects.

**Results:** In 20,064 individuals (3,353 vaccinated and 16,711 unvaccinated) hospitalized with infection due to SARS-CoV-2 between January 1^st^, 2021 and January 5^th^, 2022, vaccination with 1, 2, or 3 doses significantly reduced the risk of ICU admission and death. An inverse dose-response relationship was observed between vaccine doses received and both outcomes (adjusted odds ratio (aOR) for ICU admission per additional dose: 0.66, 95% CI 0.62 to 0.71; aOR for death per additional dose: 0.78, 95% CI 0.72 to 0.84). The reduction in risk was greater for ICU admission than for death (P for heterogeneity <0.05), but no significant differences in risk were seen based on infecting variant of concern (VOC).

**Interpretation:** We identified a decrease in the risk of ICU admission and death in vaccinated individuals compared to unvaccinated, time-matched controls, even when vaccines failed to prevent infection sufficiently severe to cause hospitalization. Even with diminished efficacy of vaccines against infection with novel VOCs, vaccines remain an important tool for reduction of ICU admission and mortality.

## Introduction

The global pandemic of SARS-CoV-2 has sickened hundreds of millions of people, and killed millions (1). The rapid development of safe and effective vaccines against the virus has been a singular scientific achievement and has likely prevented many more illnesses and deaths (2-4). However, the ongoing emergence of novel viral variants remains a challenge, with reduced protection against infection seen with the B.1.529 (Omicron) variant that emerged in autumn 2021 (5), though vaccines seem to continue preventing severe illness resulting in hospitalization, intensive care admission, and death. Indeed, the effectiveness of vaccination for prevention of severe outcomes may reflect two different effects, which may be difficult to disentangle: (i) prevention of infection, and (ii) prevention of severe outcomes even when vaccination does not prevent infection. Studies may be further complicated by factors such as decreased propensity to test vaccinated individuals with mild symptoms of respiratory illness, which could produce biases that result in increased apparent severity of infection following vaccination.

We have previously evaluated the effectiveness of SARS-COV-2 vaccines in the Canadian province of Ontario to prevent infection (6). However, vaccines that result in attenuation of severity, even among individuals in whom they failed to protect against infection, would be of considerable value to vaccinated individuals and to the wider community by limiting consumption of scarce critical care resources. We sought to study the impact of prior vaccination on severity of illness among individuals admitted to hospital with SARS-COV-2 in Ontario, Canada, as a study limited to hospitalized individuals should limit biases introduced by differential testing according to disease severity and vaccination status. As both the propensity to receive vaccination and the dominant viral variant of concern (VOC) has varied over the course of the pandemic in Ontario, we used a time-matched cohort to evaluate the adjusted risk of intensive care unit (ICU) admission and death in vaccinated and unvaccinated individuals, with identically timed infection, admitted to Ontario hospitals. Our primary objective was to determine whether the risks of ICU admission and death were diminished significantly by vaccination among individuals whose vaccination failed to prevent hospitalization. We also performed exploratory analyses to evaluate whether protective effects were modified by individuals’ characteristics, or by infecting viral variant.

## Methods

### Data Sources

We created a time-matched cohort of individuals who were hospitalized due to SARS-CoV-2 infection between January 1^st^, 2021 and January 5^th^, 2022. Individuals who had received one, two, or three doses of a SARS-COV-2 vaccine were included as exposed and were matched to unvaccinated individuals based on the test date for SARS-CoV-2. Each vaccinated individual was matched with up to five unvaccinated individuals, acting as controls (7). We identified vaccinated and unvaccinated SARS-CoV-2 cases in the Province’s Case and Contact Management (CCM) system, as described elsewhere (8, 9).

We included only cases with a unique “pseudo-health card number”, permitting linkage with the provincial vaccination COVaxON database (9), which provided information on dose administration, dates, and vaccines used. We considered individuals to be vaccinated 14 days or more after vaccine dose administration (10). We used date of positive testing as a surrogate for onset of infection; when individuals had a test date <14 days from a dose of vaccine, they were considered not to be protected by that dose. For example, an individual tested for infection 2 days after their second dose of vaccine would be considered single-dose vaccinated. While information on third vaccine doses was available in COVaxON, third doses were not widely available at the time of the analysis and a small number of hospitalized individuals had received a third dose of vaccine. Consequently, we evaluated individuals based on the number of vaccine doses received, and by classifying them as unvaccinated, single-dose vaccinated, double-dose vaccinated, or triple-dose vaccinated.

There have been several waves of the SARS-CoV-2 pandemic in Ontario, with different infecting variants prevalent for each. Initial waves were driven by the Wuhan variant, then waves were driven by Alpha in spring 2021, Delta in summer and autumn 2021 (11), and finally Omicron in December 2021 (12). As such, infecting viral variants in our analysis were classified as non-VOC, N501Y+ variant (including the Alpha, Beta and Gamma variants), or Delta variant, as described elsewhere (9). Individuals were considered infected with the Omicron variant (B.1.1.529) if they had been identified as such through whole genome sequencing, or if they were infected on or after November 10, 2021 with a strain with S-gene target failure or the N501Y mutation. Individuals were also considered infected with the Omicron variant if they had been classed as infected with an unknown VOC after December 13^th^, 2022. A flow diagram outlining creation of the cohort is presented in **Figure 1**.

**Figure 1:**
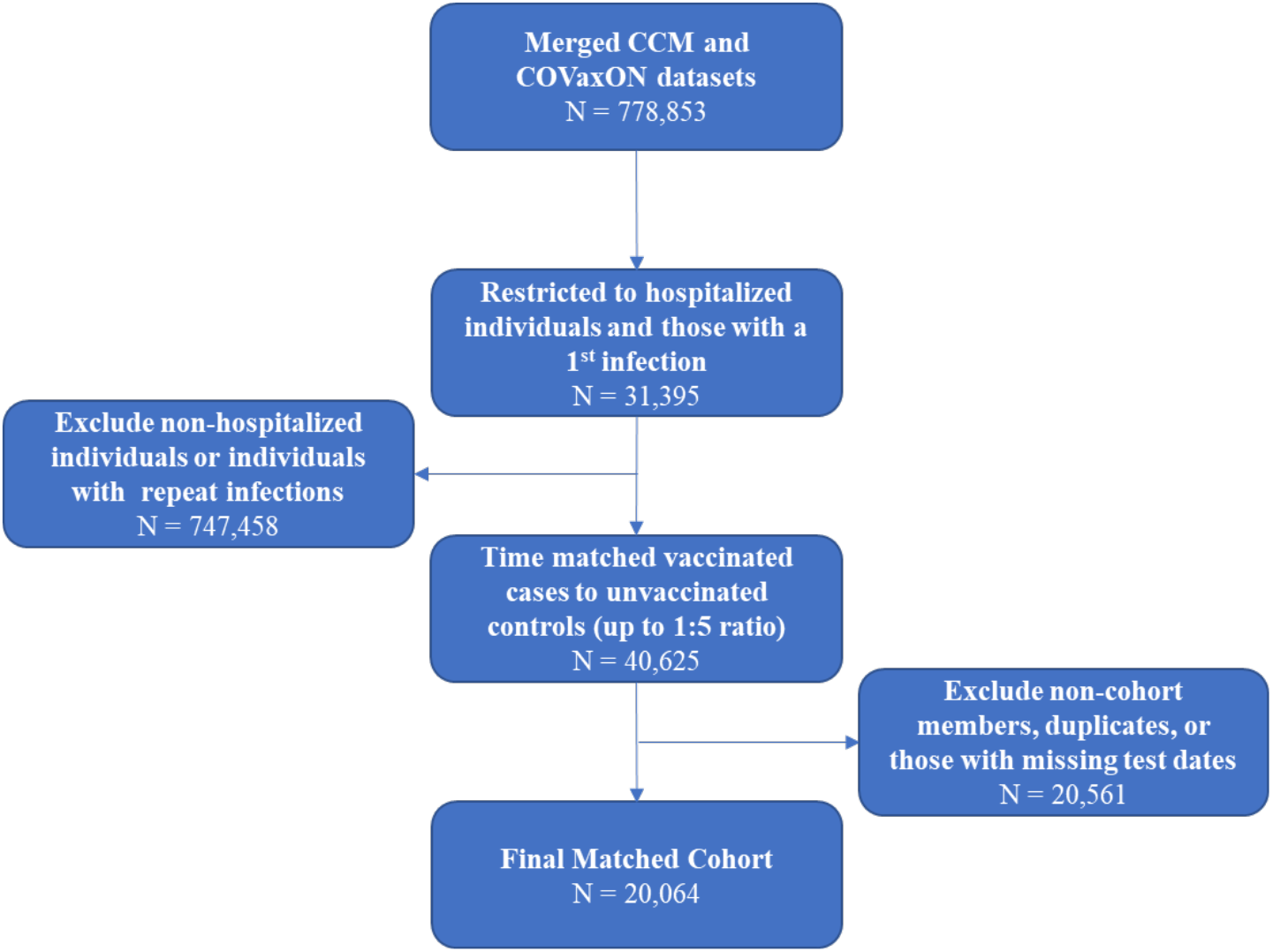
Flow diagram for the creation of the matched cohort. **NOTE**: CCM, “case and contact management system” (Ontario’s line list database); COVaxON is Ontario’s Provincial vaccination database.

### Analysis

We used our matched cohort to calculate the risk of ICU admission and death among those hospitalized due to SARS-COV-2 using conditional logistic regression models. The models were specified a priori to adjust for age category (treated as a nine-level ordinal variable), sex, healthcare worker status, long-term care residence, comorbidity, and infecting variant. Vaccine status was treated as a four-level nominal variable (zero, one, two, or three doses) where zero doses was used as the referent. Due to the rarity of deaths in healthcare workers, healthcare worker status was not included in models evaluating risk of death.

We also performed exploratory restriction analyses in which vaccinated and unvaccinated cases were limited to a single infecting variant (non-VOC, N501Y+ VOC, Delta VOC, or Omicron VOC) to evaluate modification of observed effects of vaccination of ICU admission and death by infecting variant. As conditional logistic regression models failed to converge for some of these models, we modeled the effect of vaccination using unmatched logistic regression models, with time trend modeled as a cubic trend function. Lastly, we investigated heterogeneity in the adjusted odds ratios (aORs) for ICU admission and death using meta-analytic techniques (i.e., graphically using forest plots, and statistically using the meta-analytic Q statistic). The study was conducted in accordance with the STROBE guidelines for observational research (13), and received ethics approval from the Research Ethics Board at the University of Toronto.

## Results

The final matched cohort consisted of 3,353 vaccinated individuals and 16,711 unvaccinated individuals. Most cohort members were aged 50 and over (69.47%) and most were infected with the N501Y+ VOC (33.41%). The cohort was majority male (53.69%), and 21.58% of individuals had a recorded major medical comorbidity. In univariable analyses, vaccinated and unvaccinated individuals differed significantly according to age group, residence in long-term care, comorbidity status, and infecting variant (**Table 1)**. Among vaccinated individuals, 51.12% had received one dose, 44.23% had received two doses, and only 4.65% had received 3 doses.

**Table 1:** Baseline characteristics of the matched cohort.

|  | Vaccinated | (%) | Unvaccinated Controls | (%) | Total | (%) | P-value |
| --- | --- | --- | --- | --- | --- | --- | --- |
| N | 3,353 |  | 16,711 |  | 20,064 |  |  |
| Outcome |  |  |  |  |  |  |  |
| ICU Admission | 467 | 13.93% | 3,819 | 22.85% | 4,286 | 21.36% | <0.001 |
| Death | 517 | 15.42% | 1,849 | 11.06% | 2,366 | 11.79% | <0.001 |
| Age Group |  |  |  |  |  |  |  |
| 0-9 or 10-19* | 32 | 0.95% | 1,038 | 6.21% | 1,070 | 5.33% | <0.001 |
| 20-29 | 52 | 1.55% | 905 | 5.42% | 957 | 4.77% |  |
| 30-39 | 96 | 2.86% | 1,665 | 9.96% | 1,761 | 8.78% |  |
| 40-49 | 142 | 4.24% | 2,195 | 13.14% | 2,337 | 11.65% |  |
| 50-59 | 281 | 8.38% | 3,108 | 18.60% | 3,389 | 16.89% |  |
| 60-69 | 576 | 17.18% | 3,376 | 20.20% | 3,952 | 19.70% |  |
| 70-79 | 798 | 23.80% | 2,669 | 15.97% | 3,467 | 17.28% |  |
| 80+ | 1,376 | 41.04% | 1,754 | 10.50% | 3,130 | 15.60% |  |
| Male | 1,838 | 54.82% | 8,935 | 53.47% | 10,773 | 53.69% | 0.135 |
| Healthcare Worker | 16 | 0.48% | 88 | 0.53% | 104 | 0.52% | 0.716 |
| Long-Term Care | 102 | 3.04% | 93 | 0.56% | 195 | 0.97% | <0.001 |
| Comorbidity |  |  |  |  |  |  |  |
| Any Significant Comorbidity | 1,088 | 32.45% | 3,242 | 19.40% | 4,330 | 21.58% | <0.001 |
| Asthma | 79 | 2.36% | 344 | 2.06% | 423 | 2.11% | 0.274 |
| Hematological Disease | 47 | 1.40% | 136 | 0.81% | 183 | 0.91% | 0.001 |
| Cardiac Disease | 578 | 17.24% | 1,662 | 9.95% | 2,240 | 11.16% | <0.001 |
| COPD | 142 | 4.24% | 276 | 1.65% | 418 | 2.08% | 0.628 |
| Diabetes | 344 | 10.26% | 1,110 | 6.64% | 1,454 | 7.25% | <0.001 |
| Renal Disease | 138 | 4.12% | 226 | 1.35% | 364 | 1.81% | <0.001 |
| Neurological Disease | 90 | 2.68% | 186 | 1.11% | 276 | 1.38% | <0.001 |
| Obesity | 49 | 1.46% | 244 | 1.46% | 293 | 1.46% | 0.996 |
| Liver Disease | 35 | 1.04% | 73 | 0.44% | 108 | 0.54% | <0.001 |
| Immune Compromise | 215 | 6.41% | 448 | 2.68% | 663 | 3.30% | <0.001 |
| Infecting Variant |  |  |  |  |  |  |  |
| VOC Not Detected | 69 | 2.06% | 432 | 2.59% | 501 | 2.50% | <0.001 |
| N501Y+ VOC | 1,056 | 31.49% | 5,648 | 33.80% | 6,704 | 33.41% |  |
| Delta VOC | 954 | 28.45% | 5,362 | 32.09% | 6,316 | 31.48% |  |
| Presumptive Omicron VOC | 645 | 19.24% | 2,917 | 17.46% | 404 | 2.01% |  |
| Other VOC | 49 | 1.46% | 207 | 1.24% | 256 | 1.28% |  |
| Unknown | 580 | 17.30% | 2,145 | 12.84% | 5,883 | 29.32% |  |

\*Age groups combined due to small cells ( $\leq 5$ ).
**NOTE:** COPD, chronic obstructive pulmonary disease; ICU, intensive care unit; VOC, variant of concern.

We fit two conditional logistic regression models to assess the risk of being admitted to the ICU and the risk of death in hospitalized individuals (**Table 2**). Compared to no vaccination, vaccination with a single dose of vaccine (aOR 0.57, 95% CI 0.49 to 0.66), with 2 doses (aOR 0.51, 95% CI 0.44 to 0.60), or with 3 doses (aOR 0.56, 95% CI 0.34 to 0.93) significantly reduced ICU admission risk. When vaccination status was treated as a 4-level ordinal variable, we identified a significant inverse dose-response relationship between vaccine doses received and ICU admission risk (aOR per additional dose 0.66, 95% CI 0.62 to 0.71).

**Table 2:**
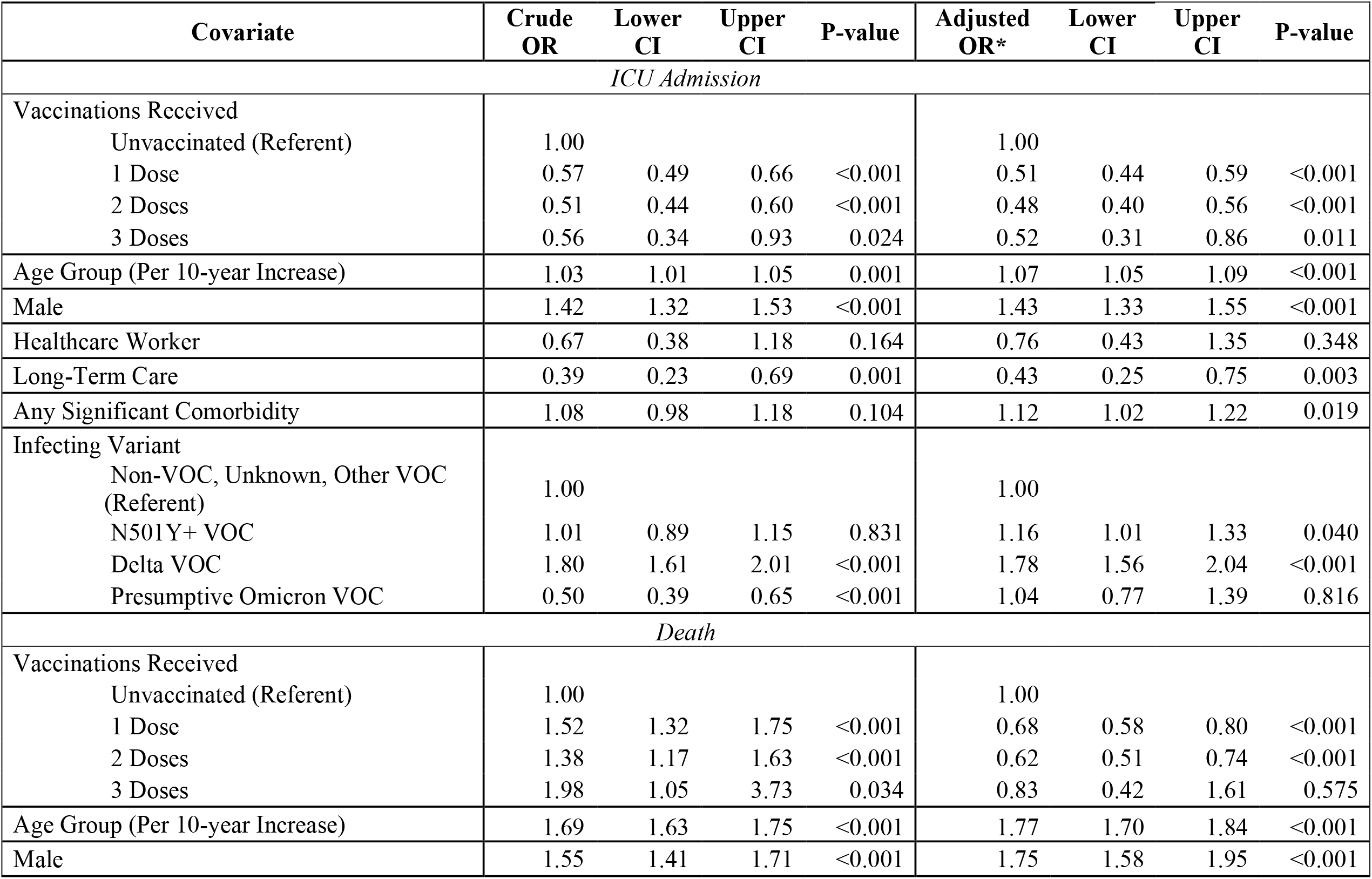
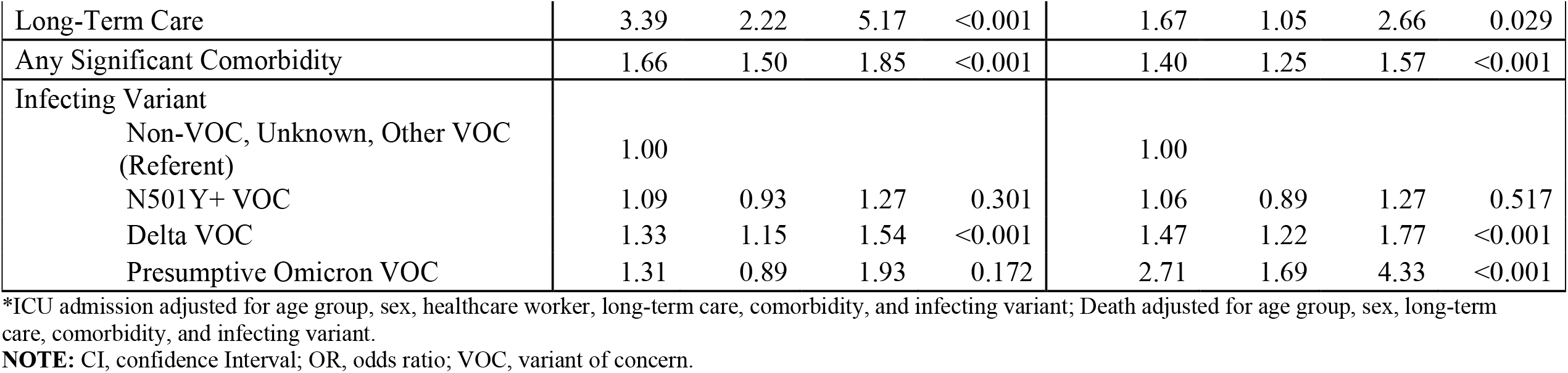
Odds ratios and 95% confidence intervals from conditional logistic regression on intensive care unit admission and death due to SARS-COV-2.

Vaccination also significantly decreased the risk of death conditional on hospitalization for SARS-COV-2 (aOR for a single vaccine dose 0.5, 95% CI 0.44 to 0.59; aOR for 2 doses 0.48, 95% CI 0.40 to 0.56; aOR for 3 doses 0.52, 95% CI 0.31 to 0.86). We identified a significant inverse dose-response relationship between the number of vaccine doses received and risk of death (aOR per additional dose 0.78, 95% CI 0.72 to 0.84).

We also performed an exploratory subgroup analysis on the odds of ICU admission and death stratified by the infecting variant. Heterogeneity in the aORs for ICU admission and death is shown in **Figure 2**. There was no significant heterogeneity between variants within each outcome. However, there was significant heterogeneity in vaccine effects based on the outcome used, with greater protection against ICU admission than death (p<0.05). Results of stratified models are presented in detail in the **Supplementary Tables**.

**Figure 2:**
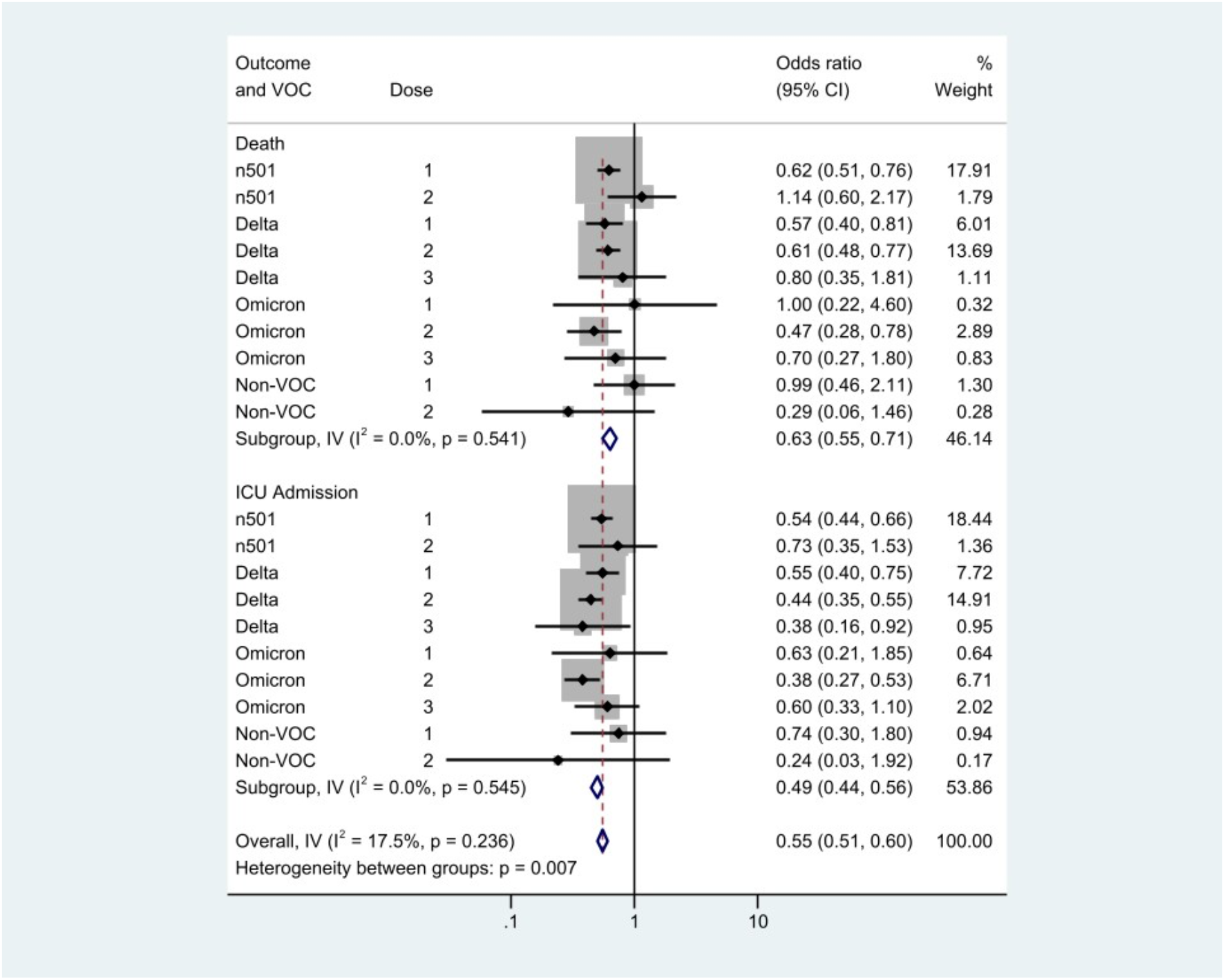
Forest plot to evaluate heterogeneity between estimates by infecting variant and outcome. The analysis is stratified by outcome, with results for death in the upper rows, and ICU admission below. **NOTE:** CI, confidence interval; ICU, intensive care unit; I-V, contribution of within-stratum variance to overall variance; n501, N501Y-positive variant; VOC, variant of concern.

## Discussion

In a cohort of vaccinated and unvaccinated individuals matched on infection timing and hospitalization with SARS-CoV-2 infection in Ontario, Canada, vaccination was associated with a decreased risk of ICU admission and death after adjustment for confounding factors such as age, sex, healthcare worker status, long-term care residence, comorbidity status, and infecting variant. A reduced risk of severe outcomes with increased number of vaccine doses received was also seen. The time-matched nature of our design suggests that our findings are unlikely to be due to changes in vaccine prioritization or dominant circulating variants over time. Restriction to hospitalized individuals makes it less likely that our findings are biased by differential testing in vaccinated and unvaccinated individuals. We postulate that this might result in confounding and selection bias, inasmuch as vaccinated individuals may be less likely to be tested for mild symptoms of infection but may be more likely to be tested overall.

The significant protective effects of vaccination against ICU admission and death were seen for both the N501Y+ and Delta variants in exploratory restriction analyses. While we did not identify significant protection against the Omicron variant in restriction analyses, this likely reflects low statistical power due to recent Omicron emergence. We found no evidence for heterogeneity in effect according to infecting variant. We did, however, identify greater protection against ICU admission than against death with vaccines. It is possible that this may relate to the protective effect seen with long-term care residence, adjusted for other covariates, but the increased risk of death associated with this factor.

Protection against infection and symptomatic infection has declined with the emergence of novel variants of concern, most notably the Omicron variant (5, 14-17). While recent data suggest that booster doses of mRNA vaccines substantially restore vaccine efficacy (16, 17), our analysis shows that prior partial vaccination can provide benefits to individuals and health systems even when vaccines fail to prevent infection, or even hospitalization, and remains an important pillar of the public health response to the SARS-CoV-2 pandemic.

The effects we demonstrate here are not unique to SARS-CoV-2 and we have previously identified similar effects with prior pneumococcal and influenza vaccination (18, 19). As with that earlier work, an important limitation here is the inability to ensure that the effects we observe are not at least in part due to residual confounding. This is a potential limitation of any cohort study; and is one that will be the focus of future work. The relatively recent emergence of the Omicron variant, and the lags associated with critical illness and death, results in the lack of statistical power to estimate Omicron-specific protections as noted above. Unfortunately, ongoing high rates of SARS-CoV-2 hospitalizations, critical illnesses, and deaths in Ontario mean that we will be able to address this potential limitation in the months ahead.

In summary, we identified a decrease in the risk of ICU admission and death in hospitalized, vaccinated individuals compared to hospitalized, unvaccinated individuals, matched for infection timing, in Ontario, Canada. Our analysis further emphasizes the critical importance of high rates of vaccination for protection of community health and reducing the impacts of SARS-CoV-2 on ICU capacity during the pandemic.

## Data Availability

Data are not publicly available. Further information on data and analyses can be obtained by contacting Prof. Fisman.

## Acknowledgements

The authors with to thank the staff at Public Health Ontario and Ontario’s public health units for collecting, sequencing, analyzing, and providing access to the data used for this analysis.

## Supplementary Tables

**Table S1:** Adjusted odds ratios and 95% confidence intervals from logistic regression on intensive care unit admission due to SARS-COV-2, stratified by infecting SARS-COV-2 variant.

| Covariate | Adjusted OR | Lower CI | Upper CI | P-value |
| --- | --- | --- | --- | --- |
| *N501Y+\** |  |  |  |  |
| Vaccinations Received |  |  |  |  |
| Unvaccinated (Referent) | 1.00 |  |  |  |
| 1 Dose | 0.54 | 0.44 | 0.66 | <0.001 |
| 2 Doses | 0.73 | 0.34 | 1.53 | 0.400 |
| 3 Doses | - | - | - | - |
| *Delta\** |  |  |  |  |
| Vaccinations Received |  |  |  |  |
| Unvaccinated (Referent) | 1.00 |  |  |  |
| 1 Dose | 0.55 | 0.41 | 0.75 | <0.001 |
| 2 Doses | 0.44 | 0.35 | 0.55 | <0.001 |
| 3 Doses | 0.38 | 0.16 | 0.92 | 0.032 |
| *Omicron\*\** |  |  |  |  |
| Vaccinations Received |  |  |  |  |
| Unvaccinated (Referent) | 1.00 |  |  |  |
| 1 Dose | 0.63 | 0.22 | 1.85 | 0.403 |
| 2 Doses | 0.38 | 0.28 | 0.53 | <0.001 |
| 3 Doses | 0.60 | 0.33 | 1.10 | 0.096 |

\*Adjusted for age group, sex, healthcare worker, long-term care, comorbidity, and time cubed.
\*\*Adjusted for age group, sex, long-term care, comorbidity, and time cubed.
**NOTE:** CI, confidence interval; OR, odds ratio; VOC, variant of concern.

**Table S2:** Adjusted odds ratios and 95% confidence intervals from logistic regression on death due to SARS-COV-2, stratified by infecting SARS-COV-2 variant.

| Covariate | Adjusted OR* | Lower CI | Upper CI | P-value |
| --- | --- | --- | --- | --- |
| *N501Y+* |  |  |  |  |
| Vaccinations Received |  |  |  |  |
| Unvaccinated (Referent) | 1.00 |  |  |  |
| 1 Dose | 0.62 | 0.51 | 0.76 | <0.001 |
| 2 Doses | 1.14 | 0.60 | 2.17 | 0.682 |
| 3 Doses | - | - | - | - |
| *Delta* |  |  |  |  |
| Vaccinations Received |  |  |  |  |
| Unvaccinated (Referent) | 1.00 |  |  |  |
| 1 Dose | 0.57 | 0.40 | 0.81 | 0.002 |
| 2 Doses | 0.61 | 0.48 | 0.77 | <0.001 |
| 3 Doses | 0.80 | 0.35 | 1.81 | 0.592 |
| *Omicron* |  |  |  |  |
| Vaccinations Received |  |  |  |  |
| Unvaccinated (Referent) | 1.00 |  |  |  |
| 1 Dose | 1.00 | 0.22 | 4.60 | 0.996 |
| 2 Doses | 0.47 | 0.29 | 0.78 | 0.003 |
| 3 Doses | 0.70 | 0.27 | 1.80 | 0.456 |

\*Adjusted for age group, sex, long-term care, comorbidity, and time cubed.
**NOTE:** CI, confidence interval; OR, odds ratio; VOC, variant of concern.

